# Development of Machine Learning Algorithms Using EEG Data to Detect the Presence of Chronic Pain

**DOI:** 10.1101/2024.09.18.24313903

**Authors:** Joseph A. Lovelace, Jonathan Miller, Skylar Jacobs, William Koppes, Frank Minella, Federica Porta, Fletcher A. White, Mark R. Hutchinson, Jonathan Miller, Skylar Jacobs, William Koppes, Frank Minella, Federica Porta, Joseph A. Lovelace

## Abstract

Chronic pain impacts more than one in five adults in the United States (US) and the costs associated with the condition amount to hundreds of billions of dollars annually. Despite the tremendous impact of chronic pain globally, the standard of care for diagnosis depends on subjective self-reporting of pain state, with no objective assessment procedure available. This study investigated the application of signal processing and machine learning to electroencephalography (EEG) data for the development of classification algorithms capable of differentiating subjects with diverse chronic pain etiologies from pain-free subjects. The study population included participants experiencing various types of chronic pain, including nociceptive, neuropathic, and mixed etiological pain conditions. Chronic pain diagnoses were based on clinical evaluation by participating physicians and adhered to the International Association for the Study of Pain (IASP) definition, requiring pain persistence for >3 months and associated functional impairment or emotional distress. Data from 186 participants were used for algorithm development, including 35 healthy controls and 151 chronic pain patients. Machine learning methodologies were applied to the data, with Elastic Net chosen as the optimal methodology.. The classifier was able to differentiate pain versus no pain subjects with an accuracy of 79.6%, sensitivity of 82.2%, and specificity of 66.7%. This study incorporates the multidimensional nature of chronic pain, ensuring that our methods and interpretations align with current clinical and research standards. This study represents a step toward integrating EEG-based biomarkers into clinical workflows for chronic pain assessment, bridging the gap between subjective reporting and objective diagnostic tools.

## 1. Introduction

Chronic pain is a pervasive and debilitating condition that affects a significant proportion of the global population. Over 20% of adults in the United States experience chronic pain, leading to substantial personal and societal burdens (Yong, 2020). Chronic pain impairs individuals’ quality of life and imposes immense economic costs, including healthcare expenditures, lost productivity, and decreased workforce participation (Gaskin, 2011). Despite its prevalence and impact, the assessment and management of chronic pain remain challenging due to the subjective nature of pain perception and reporting.

The gold standard for chronic pain assessment relies heavily on patient self-reporting using scales such as the Numeric Rating Scale (NRS) or the Visual Analog Scale (VAS). While these tools are simple and easy to administer, they are inherently subjective and susceptible to various biases, including patient variability in pain expression, recall bias, and the influence of psychological factors (Herr, 2011). Moreover, specific populations, such as individuals with communication impairments or cognitive disabilities, may be unable to effectively communicate their pain, further complicating accurate pain assessment (Thibodeau, 2006)]. Consequently, the reliance on self-report measures can lead to inconsistent and unreliable pain evaluations, hindering effective pain management and treatment personalization.

In response to the limitations of subjective pain assessment, there has been a growing interest in developing objective measures of pain. Advances in neuroimaging and neurophysiological techniques have opened new avenues for quantifying pain-related brain activity (Thibodeau, 2006). Among these, electroencephalography (EEG) has emerged as a promising tool due to its non-invasive nature, high temporal resolution, and relatively low cost compared to other imaging modalities like functional Magnetic Resonance Imaging (fMRI) or Positron Emission Tomography (PET) (Levitt, 2020). EEG captures electrical activity generated by neuronal populations, providing real-time insights into brain dynamics associated with pain perception (Pinheiro, 2016). Quantitative EEG (qEEG) features represent numerical measures derived from specific patterns in brainwave activity. These features are used in this study to identify neural signatures associated with chronic pain and serve as inputs for machine learning algorithms.

The integration of machine learning (ML) with EEG data analysis has revolutionized the potential for objective pain assessment. ML algorithms are capable of identifying complex patterns and features within EEG signals that may correlate with pain states, enabling the development of classification models that can distinguish between pain and no-pain conditions with considerable accuracy (Mussigmann, 2022). Recent studies have demonstrated the feasibility of using ML-enhanced EEG to classify chronic pain states, achieving promising levels of accuracy, sensitivity, and specificity (Levitt, 2020). These advancements suggest that EEG-based ML classifiers could serve as valuable tools in clinical settings, providing clinicians with objective metrics to complement traditional pain assessments.

Despite the promising developments, significant gaps remain in the translation of EEG and ML technologies into practical clinical applications for pain assessment. Current research often involves small sample sizes, limiting the generalizability of findings, and there is a lack of standardized protocols for EEG data collection and processing tailored specifically to pain assessment. Additionally, the heterogeneity of chronic pain conditions poses challenges in developing classifiers that can accurately capture the diverse neural signatures associated with different pain etiologies.

The present study addresses these gaps by developing a robust EEG-based ML classifier to objectively distinguish chronic pain patients from pain-free individuals. By utilizing a large and diverse sample and implementing rigorous data processing and feature selection methodologies, this research seeks to enhance the reliability and validity of EEG-based pain assessment tools. The classifier’s performance, evaluated through comprehensive metrics including accuracy, sensitivity, specificity, and area under the curve (AUC), underscores its potential utility in clinical practice. Our successful implementation of EEG-based ML classifiers for pain assessment holds significant implications for personalized pain management. Objective pain metrics can facilitate more accurate diagnosis, monitor treatment efficacy, and tailor interventions to individual patient profiles, ultimately improving patient outcomes. Furthermore, integrating such technologies into clinical workflows could reduce the reliance on subjective assessments, enhancing the consistency and objectivity of pain evaluations across diverse patient populations.

## 2. Methods

### 2.1 Ethics

Approval was obtained from Advarra Institutional Review Board (study number Pro00042433) on March 12, 2020, and subsequently registered the study on www.clinicaltrials.gov (NCT04585451). Written informed consent was obtained from all participants prior to inclusion in the study.

### 2.2 Inclusion/Exclusion Criteria

Male and female participants ages 18-80 were included in the study, and pain subjects were required to meet the IASP definition of chronic pain. Subjects with neurological disorders or other conditions affecting neurological activity were excluded from the study. Additionally, patients who may have had a reason to misrepresent their pain (e.g., patients on workers compensation) were also excluded.

Study enrollment involved recruiting and screening potential participants from the Comprehensive Pain and Wellness Center (New York City, New York), Manhattan Restorative Health Sciences (New York City and New Hyde Park, New York), and Panorama Orthopedics & Spine Center (Golden, Colorado). The study was advertised at the various institutions via posted flyers and through solicitation by the study investigators (and their respective staff) at the institutions.

In total, 334 patients were recruited for the study who met eligibility criteria and provided informed consent. 308 participants fully completed the protocol including 54 healthy controls and 254 chronic pain patients. The chronic pain patient cohort included various indications as the primary diagnosis; 150 patients with back pain, 125 patients with joint pain, 87 patients with various other sources of pain, and 14 patients for whom a primary pain source was not indicated.

Of the 308 who were initially considered eligible for analysis, eight subjects were excluded due to non- conformity with expectations underlying the recruitment strategy. Three healthy controls reported being in pain at the time of the exam (not due to the exam itself) and five chronic pain subjects reported no pain at the time of the exam. This represented a confound in the expected “in pain” versus “not in pain” associated with the control versus pain groups, and thus these eight subjects were not considered in the analysis.

Twenty-one subjects did not successfully complete EEG processing, as delineated by the quality assurance software described in 2.4.2. These failures were largely due to anomalous electrode activity not detected at the time of recording. Electrode bridging, which causes multiple electrodes to read identical values, and single electrodes remaining at a fixed voltage for a short period of time (less than 2.5 seconds) were the primary causes.

### 2.3 Study Protocol

Clinical information relating to subjects’ pain history and functional impairment was collected using standardized instruments. Characterization of subjects’ mental state was performed using the PROMIS instruments for depression and anxiety.

For the study, EEG collection was conducted using the Zeto WR-19 wireless headset which has a dry electrode array with the specifications shown in Table 1. Electrode locations were in accordance with the international 10-20 system of electrode placement (Figure 1). Linked mastoids were used as a reference.

**Figure 1:**
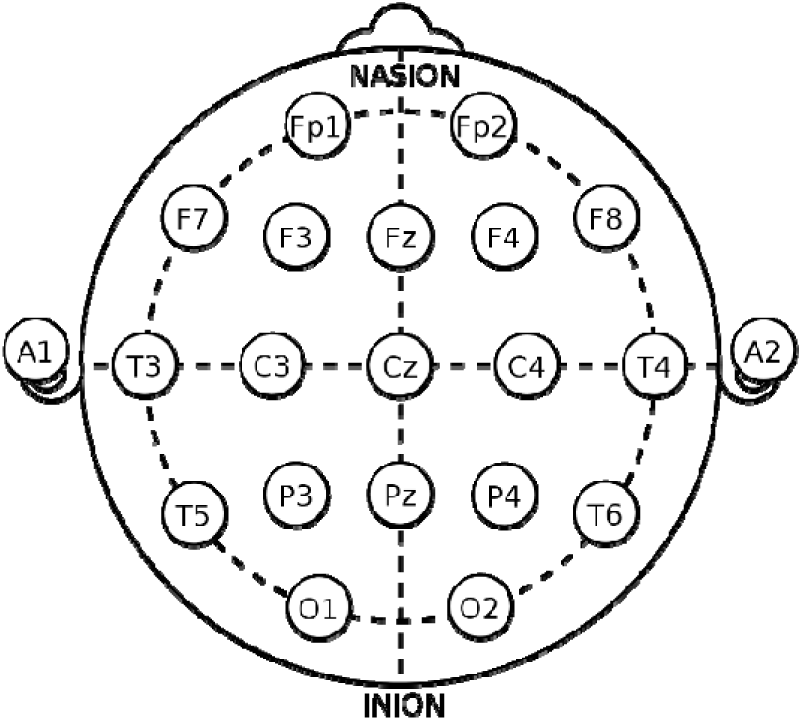
19 Channel EEG Montage showing electrodes locations in accordance with the international 10-20 System.

Fifteen minutes of eyes closed resting state EEG data were collected. Collection began after the electrode array was placed on the subject’s head and all electrode impedances were verified to be within the manufacturer’s defined acceptable range. Subjects were instructed to sit as still as possible and relax for 15 mins while the recording took place. Technicians monitored the subjects and intervened if the subject was moving excessively or began to enter a sleep state (i.e., exhibited head nodding).

### 2.4 EEG Data Processing

Following the 15 minutes of EEG data collection, the data was sent to Amazon Web Services, where it was processed by a series of software modules, developed by PainQx, to refine the data to be used for classifier development (Figure 2). The PainQx Processing Pipeline was developed with commercial intent to clean, annotate, and extract features from EEG data for the purpose of machine learning-enabled algorithm development. EEG data was supplied to the pipeline a standard European Data Format (EDF) file. EDF is a standardized way of assembling EEG signal information along with needed recording information such as sample rate.

**Figure 2:**
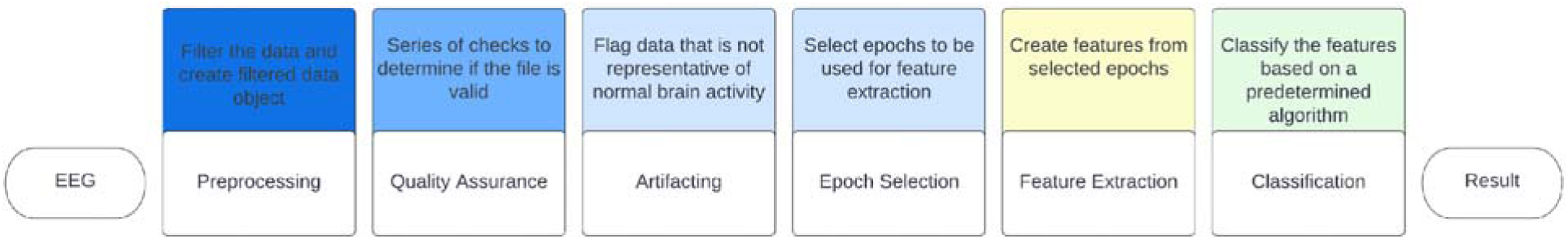
PainQx Processing Pipeline Diagram

**Figure 3.**
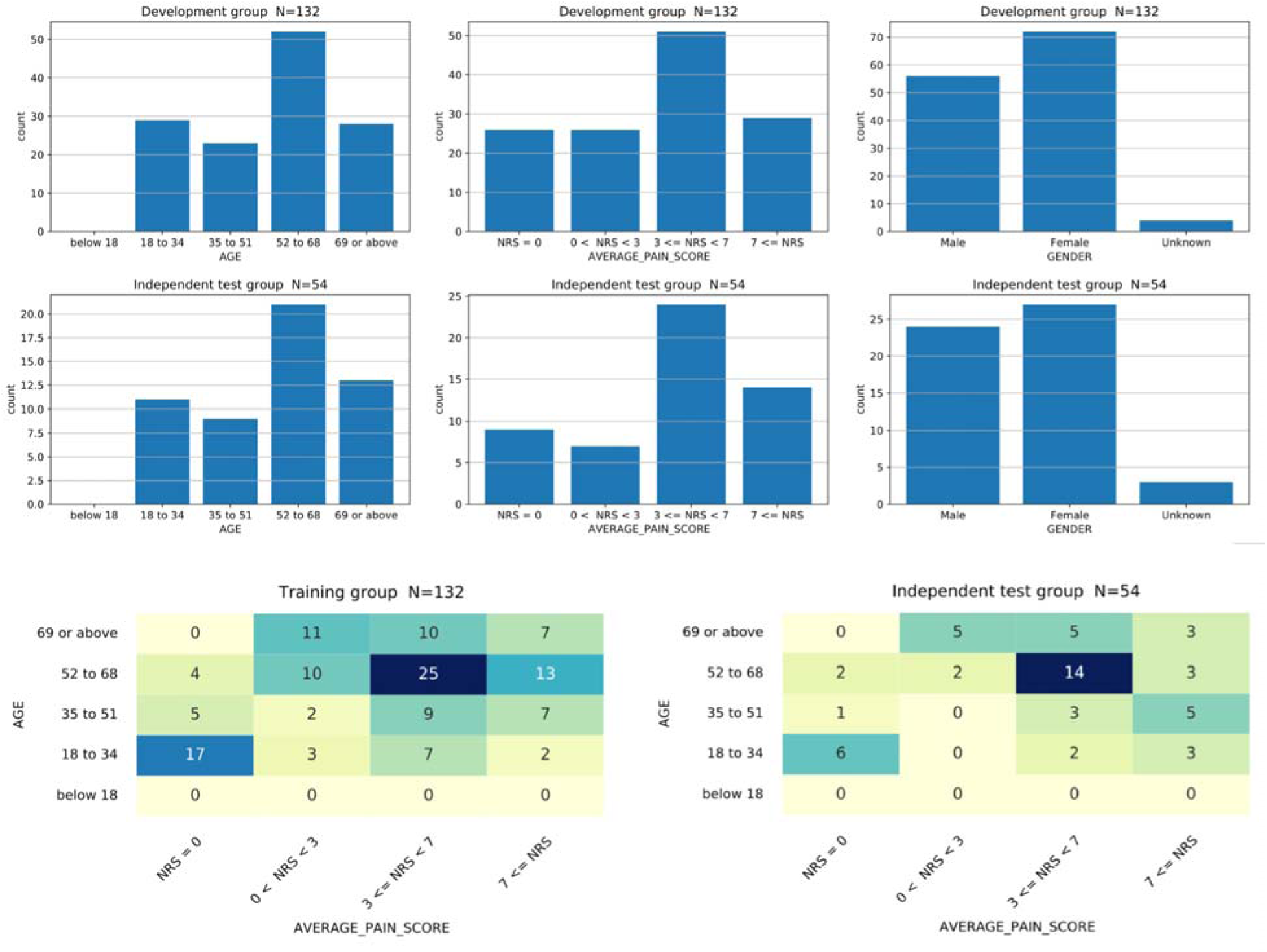
Histograms show the distribution of age, pain intensity, and gender. Heatmaps show joint distribution of age and pai intensity in the development (TT) dataset and the independent (HO) test dataset. Subjects were assigned randomly into thes groups using stratification to ensure these distributions are similar. HO recordings were set aside and processed only after classifier development was completed using only TT.

#### 2.4.1 Pre-processing

The first module in the pipeline filtered the raw EEG data by applying a second-order Butterworth bandpass filter from 1-80 Hz. An Infinite Impulse Response notch filter was also used at 60 Hz to remove powerline noise.

#### 2.4.2 EEG Data Quality Assurance

The second module in the pipeline assessed the quality of the EDF file containing the EEG data to ensure the file passed a series set of quality checks (Table 2). If any check failed, the data was deemed unusable. The initial quality assurance check ensured the data files had a header and a body. The header was first checked to verify it was readable, consisted of data obtained from standard 10-20 electrode locations, and that frequency range, and dynamic range aligned with expected values. Subsequent checks operated on the body of the EDF which contained the EEG time series data. The data resolution (least significant bit), apparent sampling rate (derived from timestamps), and power spectral density were all checked against acceptable ranges. Individual electrodes were checked to ensure good contact quality throughout the recording, and variability was compared to all other electrodes. If more than 80% of the recording was deemed to be contaminated by artifact, the file was rejected. Only if all checks passed was the file advanced to the artifactor stage.

**Table 2:**
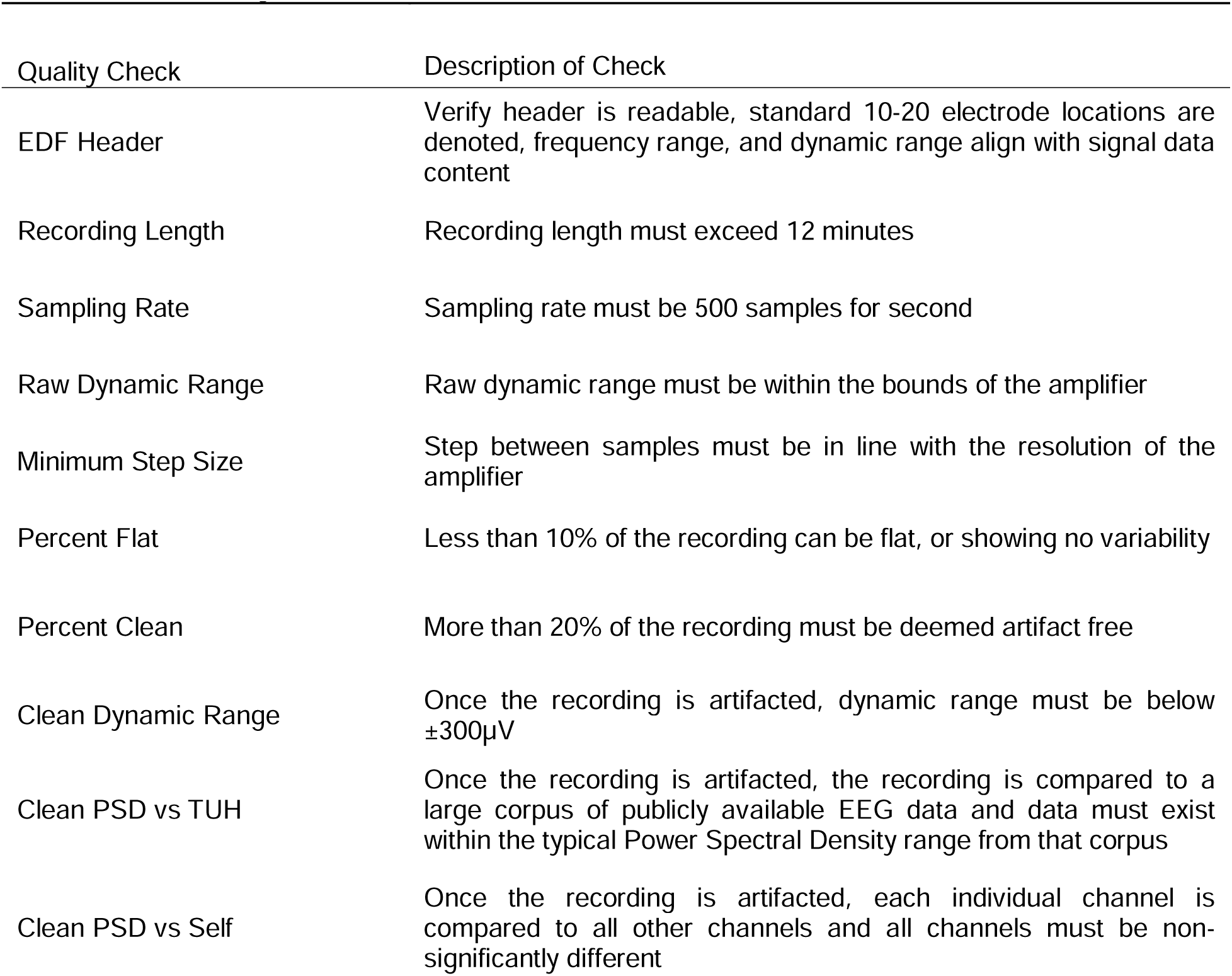
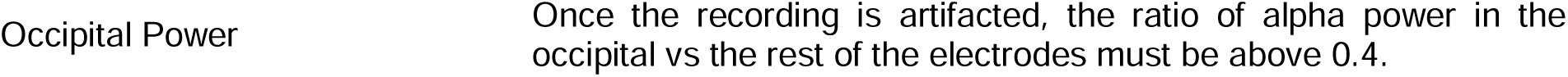
Table of Quality Control Checks.

#### 2.4.3 Epoch Creation

The full EEG recording was divided into segments referred to as “epochs,” each 2.5 seconds long. With a sample rate of 500 samples/second, each epoch contained 1250 samples (2.5 x 500) across 19 electrodes. Various epoch lengths were tested during the algorithm development process, with the previously mentioned specifications resulting in the optimal performance during cross-validation.

#### 2.4.4 Artifact Detection and Removal

The automatic artifactor software flagged any distortions in the EEG data caused by signals not originating from the brain. The artifactor produced a list of detected artifacts (Table 3), each of which was tagged with the onset time and duration in the recording. Each artifact caused an interval of EEG to be excluded. Epochs were down selected for further processing based on artifactor results with any epoch overlapping in time with an artifact being excluded from further analysis.

**Table 3:** Table of Artifact types.

| Artifact | Description |
| --- | --- |
| Impulse | Large excursion in amplitude, usually represented by a spike in the signal on one or more electrodes |
| Electromyographic | ~30 Hz noise resulting from muscle activity on one or more electrodes |
| Vertical Eye Movement | Artifact resulting from the eye moving vertically because of blinking or eye flutter, localized to Fp1, Fp2, F7 and F8 |
| Significantly Low Activity Signal | Signal is constant or near constant on one or more electrodes |

#### 2.4.5 Epoch Ranking and Selection

The epochs that remained after artifact removal were then down-selected a second time based on covariance, a method derived from the work of Congedo, Barachant et al. (Congedo, 2017). For each epoch, the covariance matrix was generated, which is a 19 -by- 19 square diagonal positive-definite matrix (given the 19 electrodes used for data acquisition). The resulting array of matrices was then analyzed to determine a single covariance matrix representing the centroid covariance of all epochs, similar to finding the centroid of a cloud of points. All epochs with distance beyond the starting threshold of 6 were excluded. The process was then iterated: 1) find centroid of epochs under analysis, 2) calculate distance and remove epochs from analysis if their distance is beyond the threshold, 3) repeat. For each iteration, the threshold was reduced by 0.95. The intent was to determine the covariance cloud which represented the “normal” covariance for the recording. Iteration was stopped when a targeted percent of epochs remained in analysis, or when the remaining epoch cloud was tight (i.e., there were no more discarded epochs beyond the distance threshold). This resulted in a single centroid derived from the epochs remaining in analysis. Finally, distance was calculated to this centroid for all epochs, including those discarded from analysis, with the final calculation performed using Riemannian distance. The rational was that Euclidean distance was much less computationally complex, so it was used in the iterative process, while Riemannian distance was a superior choice for final evaluation, justified by the fact that covariance matrices are always positive definite, thus occupying a subspace of all possible matrices, and similarity between them is reflected by distance along the subspace manifold, calculated as Riemannian distance. This method is particularly suited to pain measurement as it captures the precise and reproducible neural signatures associated with chronic pain. The covariance-based approach ensures that epochs most representative of chronic pain-related brain activity are prioritized, aligning with findings in the literature that neural oscillatory changes, particularly in the alpha and beta bands, are key biomarkers for chronic pain.

#### 2.4.6 QEEG Feature Extraction

Epochs selected for further processing by the artifacting and epoch ranking modules were then passed to the feature extractor module. Over six thousand unique EEG features were calculated for each EEG recording. The “feature type” refers to the general type of mathematical analysis performed. Those analyses were carried out for all electrodes or electrode pairs as well as for each frequency band defined in table 4, which is what led to the large number of individual qEEG features shown in table 5.

**Table 4:** Descriptions of frequency bands that were used in generation of features.

| Frequency Band Name | Symbol | Frequency Range (Hz) |
| --- | --- | --- |
| Theta | T | 3.5 – 7.5 |
| Low Alpha | A1 | 7.5 – 10.0 |
| Alpha | A | 7.5 – 12.5 |
| High Alpha | A2 | 10.0 – 12.0 |
| Beta | B | 12.5 – 25.0 |
| High Beta | B2 | 25.0 – 35.0 |
| Gamma | G | 35.0 – 50.0 |
| Full Lower Spectrum | S | 1.5 – 25.0 |

**Table 5:**
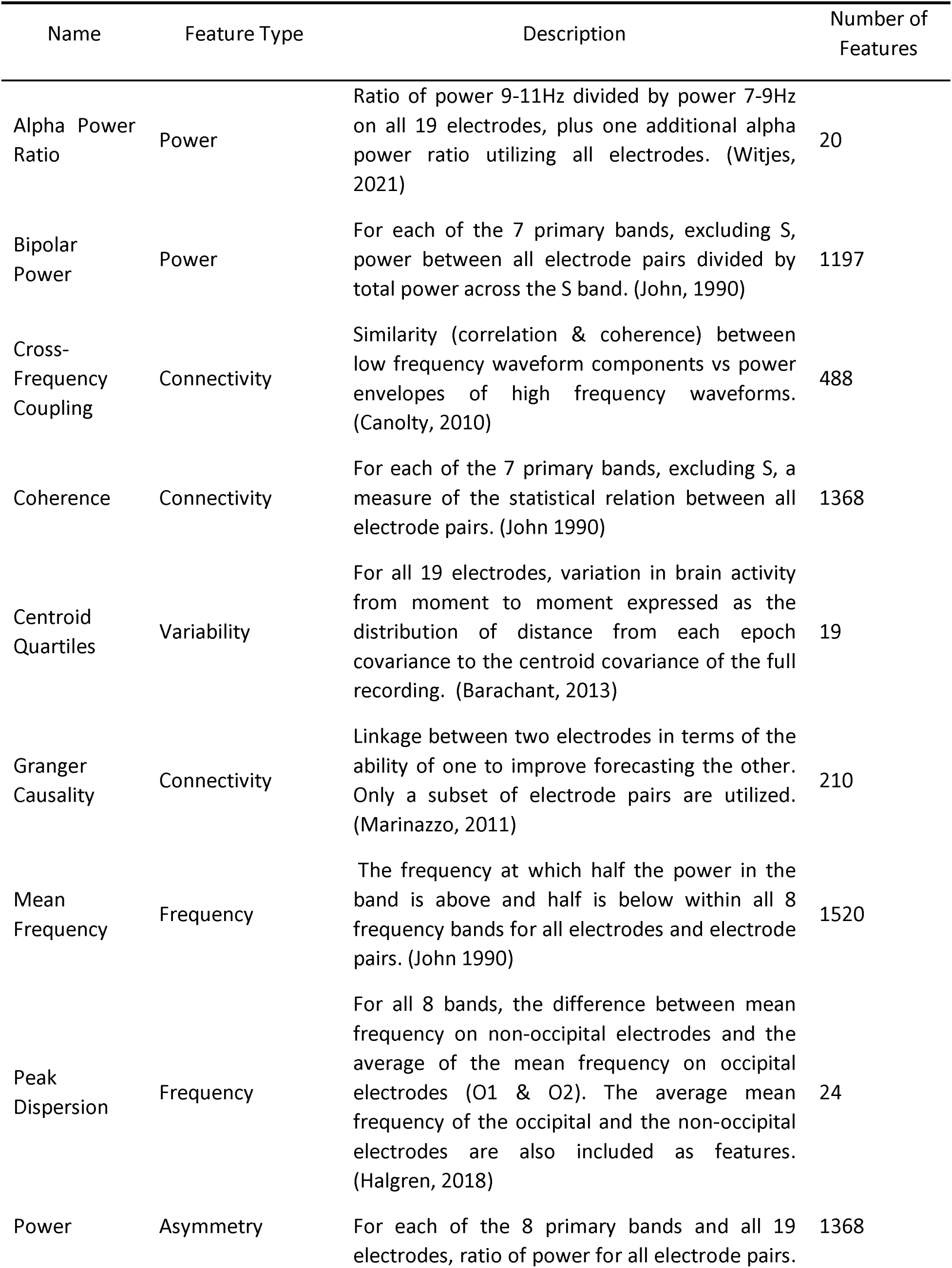
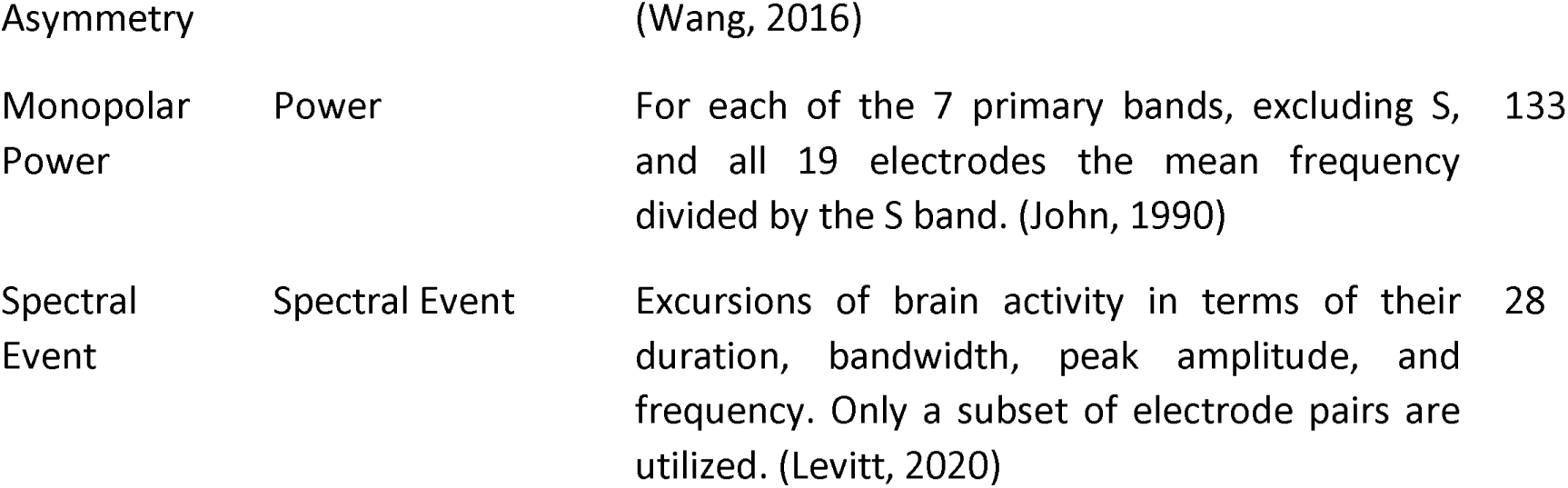
QEEG Features Calculated.

#### 2.4.7 Implications of Age on QEEG Features

Nearly all qEEG features have an expected trend with age. This was accounted for by converting features into Z-scores that incorporate age-expectation using a separate normative dataset from the Brain Research Laboratories of NYU School of Medicine which included 92 subjects with representation across ages 19 to 81. This norming dataset was used to fit a trend line for each feature. Subjects in this normative dataset did not have any evidence of clinically significant pain, depression, or anxiety, other than one individual with moderate anxiety. In each case, age dependence was assumed to be linear with log of age.

#### 2.4.8 Classifier Development

The calculated qEEG features and participant reported NRS scores were used as input to AI/ML based classifier development methodologies with the objective of developing algorithms for the characterization of chronic pain. ML best practices were utilized in the development of a pain versus no-pain binary classification algorithm, to reduce the possibility of overtraining in which results are good on the training dataset, but poor in the broad population. The approach to mitigate overtraining included rigor in the cross validation so that test folds are truly isolated from the training folds, i.e. no information leakage was allowed to occur. In addition, ML results were compared to domain knowledge to identify solutions which do not align with relationships reported in literature, potentially representing something about the dataset that may be different from the broader population. Finally, 30% of the data was randomly set aside as an independent set of data to confirm the findings of the cross validation, and this 30% “hold out” group was only processed once at the end of development, producing a clean estimate of performance on the broad population.

### 2.5 Analysis of Classifier Performance

#### 2.5.1 Train/Test and Hold Out Data Sets

Algorithms were developed on a Train/Test (TT) dataset (70% of the data), with performance evaluated on a Hold Out (HO) dataset (30% of the data) to check for potential overtraining of the algorithm. The performance of the Pain / No-pain classifier was analyzed using a variety of metrics. Performance was evaluated for both the TT and HO datasets. The TT performance was derived from 20 repetitions of 10- fold cross-validation, in which the TT set is partitioned at random into a Train interval and Test interval and results are aggregated, whereas the HO performance was a single application of the TT-developed classifier applied to the HO dataset.

#### 2.5.2 Discriminant Score and Receiver Operating Characteristics Plots

The pain versus no-pain algorithm combined weighted qEEG values to produce a discriminant score. One method for visually assessing how well a binary classifier can separate two classes is a histogram of the discriminant scores for each of the two classes, which was generated as part of the study. Another method for visualizing the performance of a binary classifier, which was also utilized as part of the study, is the Receiver Operating Characteristics (ROC) curve. The ROC curve is a performance measurement for the classifier at all possible settings of the operating point, and so it represents an unbiased perspective on performance.

#### 2.5.3 Area Under the Curve (AUC)

The Area Under the Curve (AUC) metric is derived from the ROC curve and represents the degree or measure of separability. It tells how much the model is capable of distinguishing between classes. The higher the AUC, the better the model is at predicting negative (no-pain) classes as negative and positive (pain) classes as positive. By analogy, the higher the AUC, the better the model is at distinguishing between patients with the disease/condition versus without the disease/condition.

#### 2.5.4 Operating Point Selection

To generate specific performance metrics based on the AUC – ROC curve, an operating point must be established which determines the discriminant score cut point that defines which cases are classified as positive (pain) versus negative (no-pain). The selection of an operating point is often made based on a risk/benefit analysis for a specific application of the classifier, where false positive versus false negative results may have very different risks associated with them. Since a pain vs no-pain classifier may see several different applications, the study utilized an operating point that minimized the total number of false classification results.

#### 2.5.5 Performance Metric Calculations Using Study Data

Having chosen an operating point, the application of the classification algorithm yields a specific set of true positive (TP), true negative (TN), false positive (FP) and false negative (FN) results/counts. Those counts are often displayed in a 2x2 table referred to as a confusion matrix. Utilizing the data from the confusion matrix, a variety of performance measures were calculated, each providing a different perspective on the behavior of the algorithm. Definitions for the metrics calculated as part of the study include the following:

Accuracy: Percent of correctly classified cases involving both pain and no-pain classifications

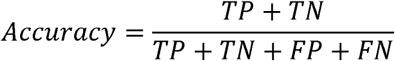

Sensitivity: Percent of correctly classified pain cases

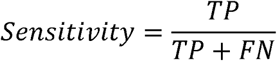

Specificity: Percentage of correctly classified no-pain cases

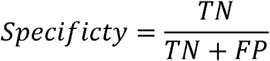

Positive Predictive Value: Percent of subjects classified as in pain who are in pain

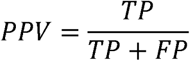

Negative Predictive Value: Percent of subjects classified as no-pain who are not in pain

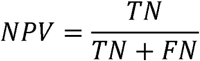

#### 2.5.6 Extended Performance Metric Calculations Using Study Data and Prevalence Models

Several of the performance metrics calculated, such as sensitivity and specificity, involve only one class, and so they are not impacted by the distribution of pain versus no-pain cases in the study population. Other metrics, such as Positive Predictive Value (PPV) and Negative Predictive Value (NPV), are impacted by changes in the ratio of positive (pain) to negative (no-pain) cases in the population under test. PPV and NPV often play an important role in the assessment of benefit versus risk associated with the use of a classifier for a specific purpose in a specific population. For this study, the goal of subject recruitment was to achieve a reasonable balance between classes (pain and no-pain) to optimize the ML process. While that goal was achieved, the resulting study population was not representative of any of the populations that might be targeted for pain / no-pain assessment in the “real world.” Therefore, in order to derive relevant PPV and NPV metrics, the Sensitivity and Specificity measures defined above were used to derive new confusion matrix values based on two “real world” prevalence models for pain versus no-pain. The first prevalence model was based on the incidence of chronic pain across adults in the U.S., which is reported to be approximately 20% (with 80% not in pain) (Yong, 2022). The second prevalence model targeted the population of patients being seen in U.S. pain clinics. That model was developed based on discussions with pain clinicians and assumed that 95% of patients being seen are experiencing pain (with 5% not in pain). Application of the sensitivity and specificity measures across each given population resulted in a new confusion matrix consisting of derived values referred to as TP, TN, FP, and FN . Those new sets of counts were used to derive PPV and NPV numbers specific to the defined population.

Positive Predictive Value (for population x):

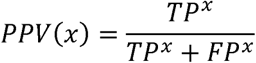

Negative Predictive Value (for population x):

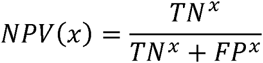

## 3. Results

### 3.1 Data Set Generation Results

The data from 132 subjects (70% of the data) was used for algorithm development and the remaining 54 (30% of the data) was used as an independent test set. Participants were assigned to either group randomly, but stratified so that distributions of gender, age, and pain intensity were similar. (Figure 4)

**Figure 4.**
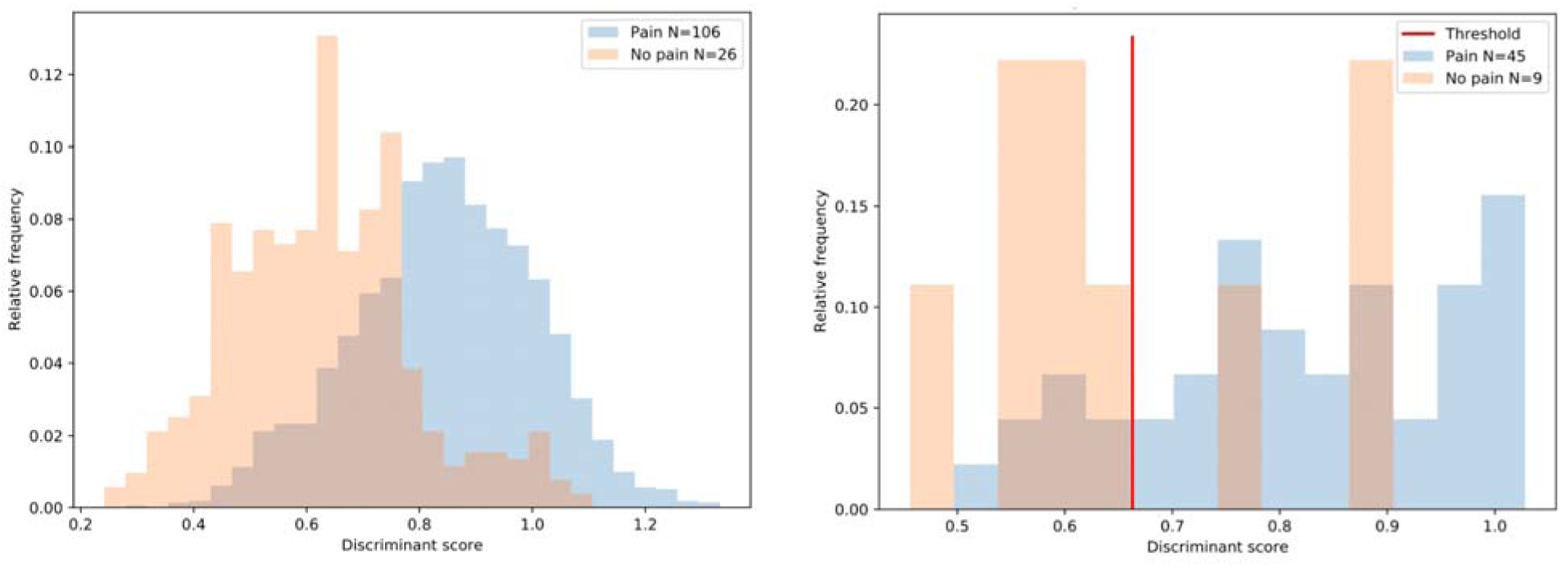
Histograms of discriminant scores for the development dataset (left panel) and the independent test set (right panel). The left panel shows test scores from the cross validation in which N=132 subjects each contributed 20 scores reflecting 2 repetitions of 10-fold cross validation. The right panel shows a single application of the classifier to an independent test set of N=54, which was processed only once.

### 3.2 Classification Algorithm Development Results

Given that the optimal ML methodology for a given classification problem can differ depending on the underlying phenomenon driving the differences between the defined classes, a variety of ML tools were applied including Gradient Boosted Trees, Nearest Neighbor Models, Logistic Regression Classifiers, Support Vector Machines, and Elastic Net Classifiers, each of which included many settings which were explored in detail. For example, Support Vector Machines were implemented with four different kernel types: Radial Basis Function, Polynomial, Sigmoid, and linear. From this work, we selected Elastic Net (ENET) (Hastie, 2009) as our preferred tool. Elastic Net was selected for its ability to handle high-dimensional data while balancing feature selection and multicollinearity, which is particularly important when working with qEEG data. The model’s sparse representation facilitates interpretability, allowing the identification of clinically meaningful biomarkers relevant to chronic pain. Cross-validation results were the dominant criteria for selection, but we also inspected details of the solutions to assess the contribution of different inputs (features) and whether highly weighted qEEG features met with expectations based on the relationships between neurological changes and chronic pain reported in the literature.

Elastic Net is a generalized method that encompasses two well-established methods: Ridge Regression and LASSO. Ridge Regression seeks to minimize the total magnitude of assigned weights (contributions) across all inputs (features), and LASSO seeks to minimize the total number of non-zero weights. ENET allows a smooth transition between the two so the objective function can reflect both goals to better fit a wide range of problems. The study utilized an L1_ratio of 0.5, meaning the solution was halfway between pure Ridge Regression and pure LASSO, and an Alpha parameter of 0.1, meaning 10% of the objective function was driven by driven by minimizing the number of features receiving non-zero weights, and the remaining 90% was driven by model match to data (i.e., least squared error).

### 3.3 Discriminant Score Results

Results for pain and no pain are shown in Figure 5. The left panel shows histograms of discriminant scores from the Development group (N=132), which arise from two hundred repetitions of 10-fold cross validation, and the right panel shows discriminant scores for the final classifier applied once to the independent test group (N=54). Statistical analysis was performed to determine if a specific pain type (i.e. nociception, neuroplastic, or neuropathic) were misclassified more frequently. No statistical significance was found. \

**Figure 5.**
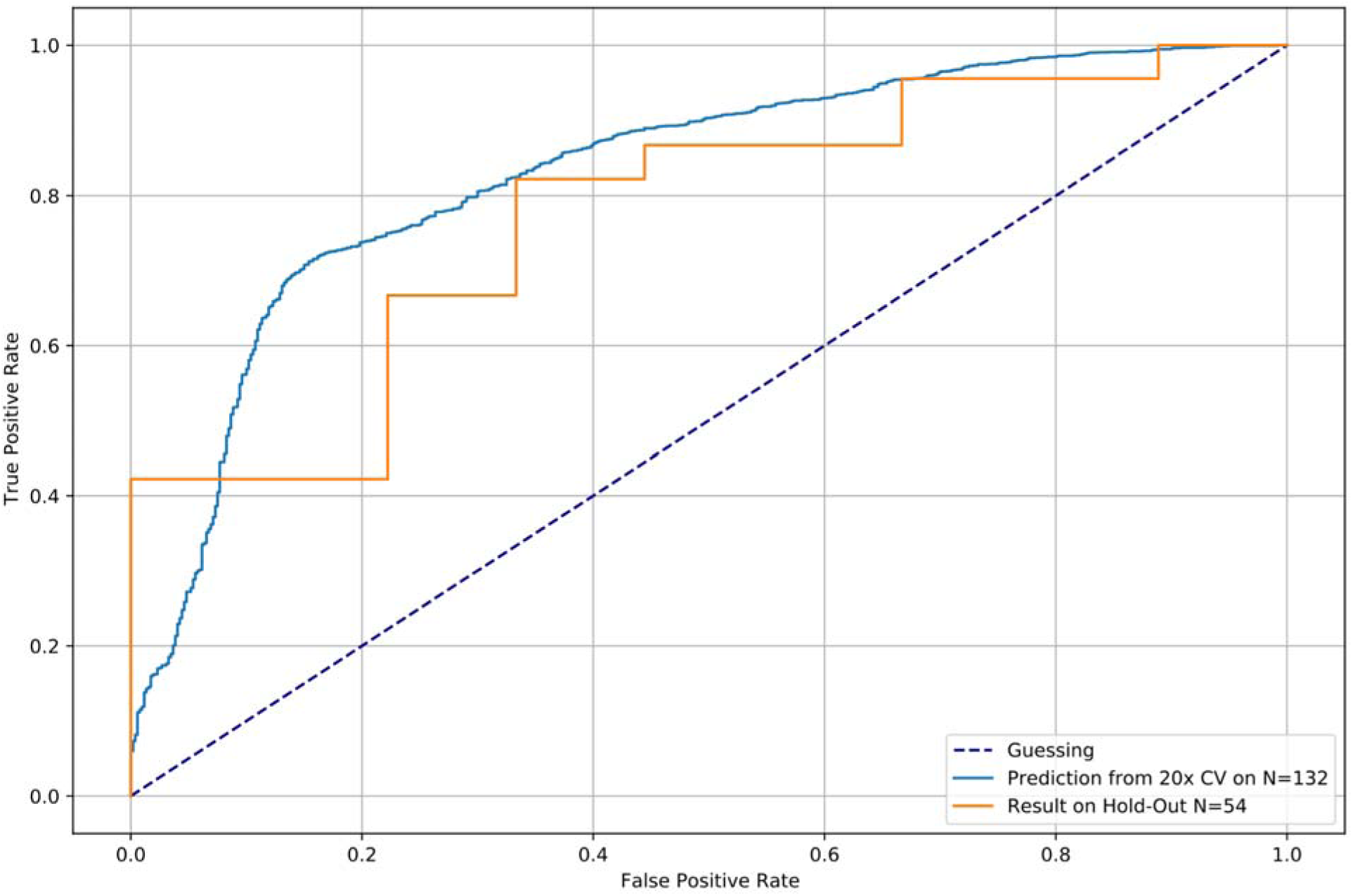
Receiver Operator Characteristic curves for the development dataset and the independent test set. The blue line is based on cross validation in which N=132 subjects each contributed 20 scores reflecting 20 repetitions of 10-fold cross validation. The orange line shows a single application of the classifier to an independent test set of N=54, which was processe only once.

### 3.4 Receiver Operating Characteristics (ROC) and Area Under the Curve (AUC) Results

The ROC curves for the TT and HO groups are shown in Figure 6. The ROC curves demonstrate the ability to classify pain versus no-pain subjects using QEEG features. The AUC calculated from the development group (TT) was 0.83, calculated as the average AUC in test subsets across 20 repetitions of 10-fold cross-validation, and AUC in the independent test group (HO) was 0.78, calculated from the single application of the final classifier trained by the development group (TT).

**Figure 6.**
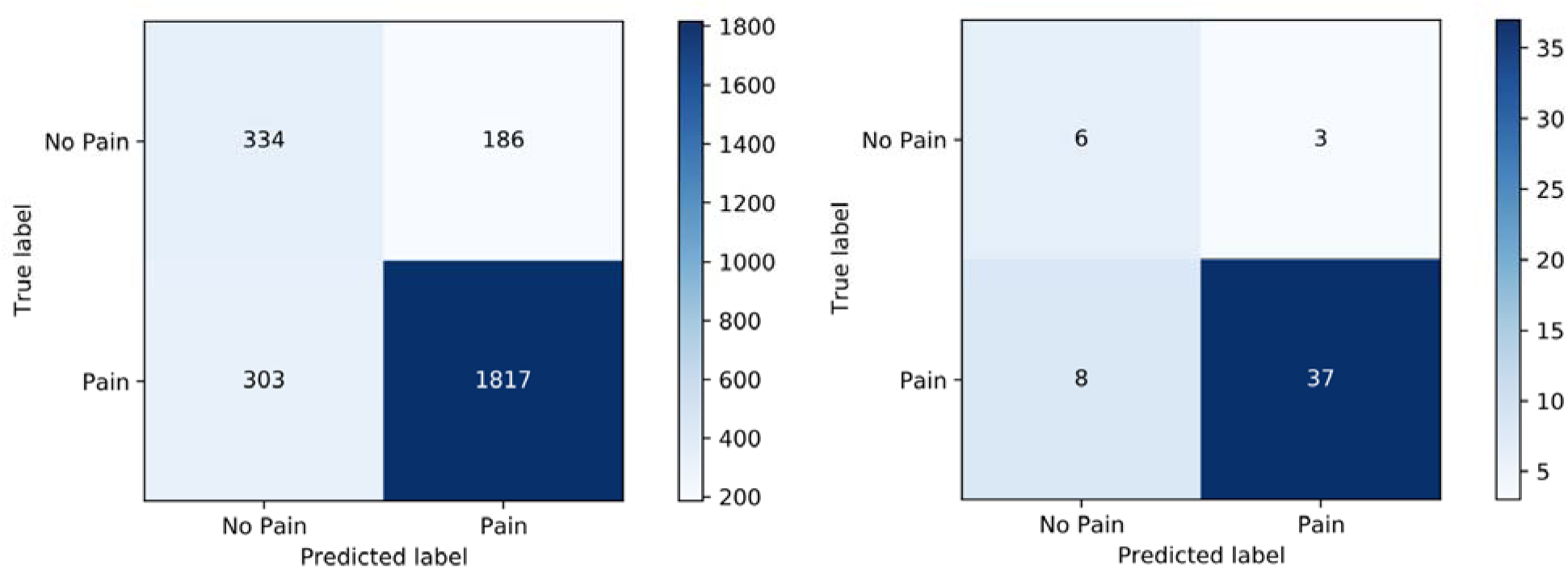
Confusion matrices show subject counts in the development dataset (left panel) and independent test (right panel). The left panel shows discriminant scores for subjects in the test interval of the cross-validation, so each subject contributes 20 values to this matrix since the CV was repeated 20 times. The right panel shows subject counts for the single application to the independent test set.

### 3.5 Confusion Matrix Results

With the operating point set to minimize total false classification results, confusion matrix results for both the TT and HO groups are illustrated in figure 7.

**Figure 7:**
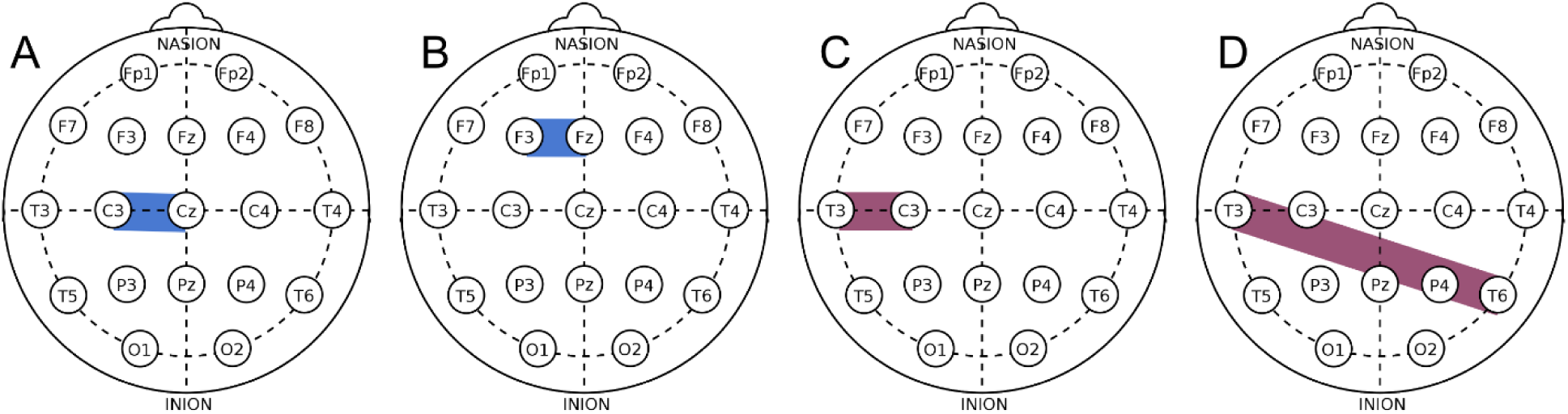
Out of the top 10 highest weighted features, two are based in Alpha band (8.5 - 12.5 Hz) shown in blue: A) Peak Alpha between C3 and Cz. B) Peak Alpha between F3 and Fz. Two others represent connectivity in different bands, shown in purple: C) Coherence between T3 and C3. D) Coherence between T3 and T6. Weights derived by machine learning show alignment with consensus in literature: Peak Alpha tends to decrease, and connectivity tends to increase with chronic pain.

### 3.6 Classification Algorithm Quality Metrics Results

Processing the data from the confusion matrices as described in the methods section of this paper. Accuracy was 0.818 for TT and 0.775 for HO. Sensitivity/specificity was 0.858/0.654 for TT and 0.822/0.667 for HO. The full set of quality metrics for both the Train/Test and Holdout groups are shown in Tables 6. Confidence intervals shown in Table 6 reflect the variability in the twenty repetitions of 10- fold cross validation.

**Table 6.** Pain / No-pain Classifier Quality Metrics

| Metric | Train / Test Dataset | Holdout (HO) Dataset |
| --- | --- | --- |
| AUC | 0.829 ± 0.016 | 0.775 |
| Accuracy | 0.815 ± 0.022 | 0.796 |
| Sensitivity | 0.857 ± 0.022 | 0.822 |
| Specificity | 0.642 ± 0.050 | 0.667 |
| NPV | 0.194 ± 0.029 | 0.165 |
| PPV | 0.978 ± 0.003 | 0.979 |

### 3.7 Extended Quality Metric Utilizing Prevalence Models Results

The study population included 151 pain cases and 35 no pain cases. Thus, the prevalence of pain in the study population was 81.2%. In order to estimate PPV and NPV for populations in which the pain versus no-pain classifier might be utilized, two prevalence models were applied as described in the methods section. Table 7 summarizes the results when the two prevalence models were applied to re-calculate PPV and NPV.

**Table 7.** PPV and NPV adjusted based on the prevalence of chronic pain in the general US adult population (20% prevalence) and a representative pain clinic population (95% prevalence)

| Metric | General US Adult Population | Outpatient Pain Clinic Population |
| --- | --- | --- |
| PPV | 0.37 | 0.99 |
| NPV | 0.95 | 0.06 |

### 3.8 QEEG Feature Selection and Weighting Results

Using ENET, specific qEEG features were selected and assigned a weight for use in the pain versus no-pain classification algorithm, with the weight assigned to a feature indicating that feature’s relevance to the classification of Pain vs No-pain. Table 8 illustrates the top 10 most highly weighted qEEG features.

**Table 8:**
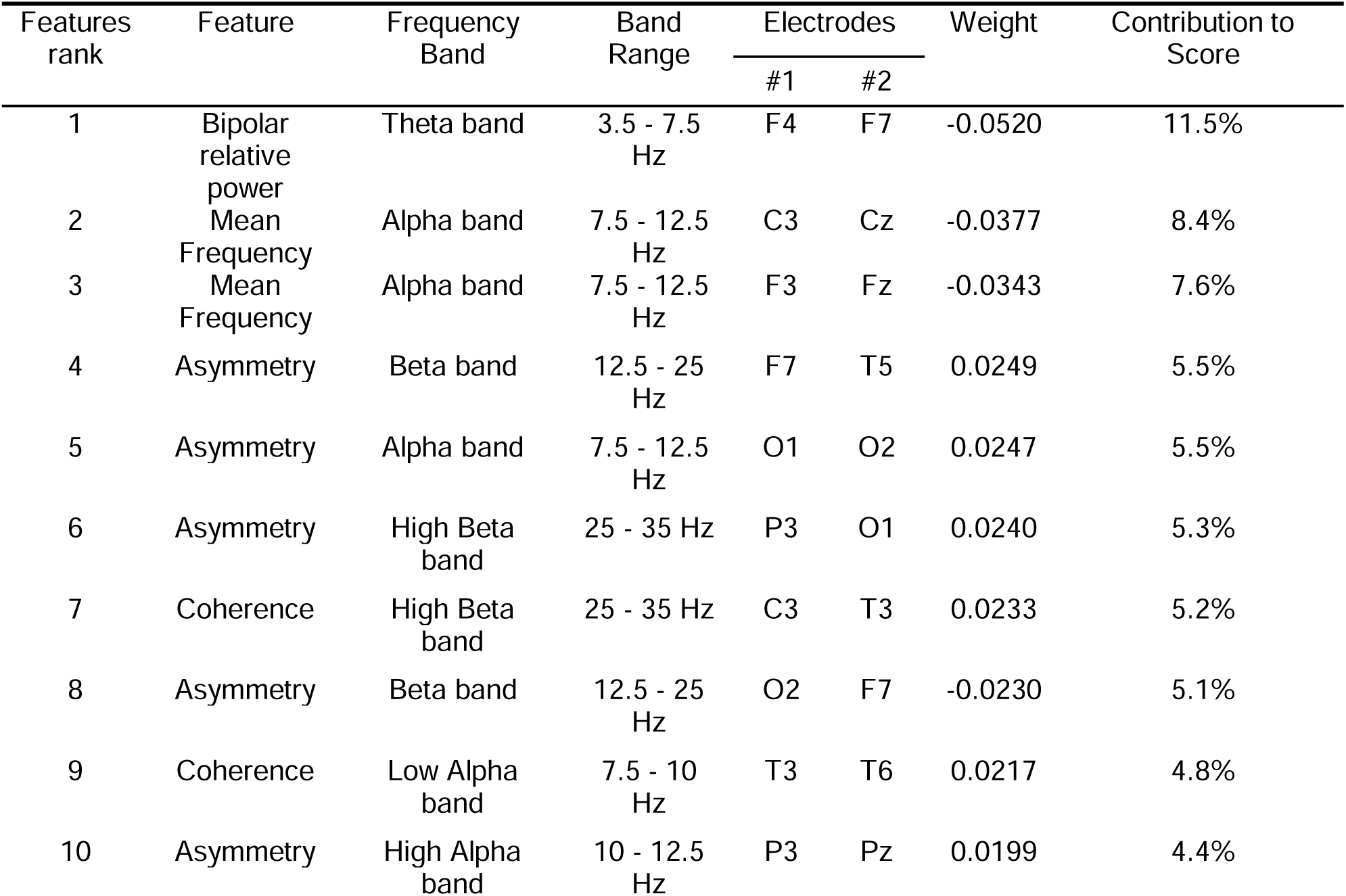
Top 10 features ranked by weight.

| Features rank | Feature | Frequency Band | Band Range | Electrodes |  | Weight | Contribution to Score |
| --- | --- | --- | --- | --- | --- | --- | --- |
|  |  |  |  | #1 | #2 |  |  |
| 1 | Bipolar relative power | Theta band | 3.5 - 7.5 Hz | F4 | F7 | -0.0520 | 11.5% |
| 2 | Mean Frequency | Alpha band | 7.5 - 12.5 Hz | C3 | Cz | -0.0377 | 8.4% |
| 3 | Mean Frequency | Alpha band | 7.5 - 12.5 Hz | F3 | Fz | -0.0343 | 7.6% |
| 4 | Asymmetry | Beta band | 12.5 - 25 Hz | F7 | T5 | 0.0249 | 5.5% |
| 5 | Asymmetry | Alpha band | 7.5 - 12.5 Hz | O1 | O2 | 0.0247 | 5.5% |
| 6 | Asymmetry | High Beta band | 25 - 35 Hz | P3 | O1 | 0.0240 | 5.3% |
| 7 | Coherence | High Beta band | 25 - 35 Hz | C3 | T3 | 0.0233 | 5.2% |
| 8 | Asymmetry | Beta band | 12.5 - 25 Hz | O2 | F7 | -0.0230 | 5.1% |
| 9 | Coherence | Low Alpha band | 7.5 - 10 Hz | T3 | T6 | 0.0217 | 4.8% |
| 10 | Asymmetry | High Alpha band | 10 - 12.5 Hz | P3 | Pz | 0.0199 | 4.4% |

The alignment with domain knowledge and published literature of 83% is largely driven by qEEG features that have strong neurophysiological relevance to chronic pain. The most influential features include bipolar relative power in the theta band (11.5%) and mean frequency in the alpha band (8.4% and 7.6%). These features, along with measures of asymmetry and coherence across alpha, beta, and gamma bands, collectively contribute to over 70% of the model’s classification score. Notably, these findings are consistent with previous studies linking alpha and beta oscillations, as well as connectivity patterns, to chronic pain states (e.g., Thibodeau, 2006; Napadow, 2010). This demonstrates the critical role of qEEG biomarkers in identifying pain-related neural signatures and underscores their importance in the classifier’s performance.

## 4. Discussion

The present study demonstrates the potential of machine learning (ML) algorithms applied to quantitative electroencephalography (qEEG) data in objectively classifying individuals experiencing chronic pain versus those who are pain-free. Achieving an accuracy of approximately 80% in both the Train/Test (TT) and Hold-Out (HO) datasets, with sensitivities exceeding 80%, these findings align with previous research indicating the viability of EEG-based biomarkers for pain assessment (Levitt, 2020; Mussigmann, 2022).

The selected qEEG features in this study, particularly those within the Alpha and Beta frequency bands, corroborate established neurological patterns associated with chronic pain. Decreased mean frequency in the Alpha band has been consistently observed in chronic pain populations, reflecting altered neural oscillations Klimesch, 1993; Mussigmann, 2022). Additionally, increased coherence in Beta bands suggests enhanced connectivity between cortical regions, which has been implicated in the maintenance and perception of chronic pain (Pinheiro, 2016; Ploner 2018. The use of Elastic Net regression to identify relevant features strengthens the reliability of the classifier by reducing overfitting and enhancing generalizability, as supported by methodological advancements in ML applications for EEG data (Zou, 2005; Hastie 2024). The consistency of classifier performance between the TT and HO datasets further validates the robustness of the selected features and the overall approach.

A unique feature identified through our analysis was a power feature in the Theta band, calculated as Bipolar Relative Power on the electrode pair F4 and F7. This feature has a negative weight in the classifier. While contemporary literature indicates that Theta oscillations tend to increase in chronic pain subjects, this is a measure of relative power that does not respond to broad changes across all electrodes and bands, but instead only indicates changes in the given electrode pair relative to all others, which is not a focus of contemporary literature and warrants further study.

The International Association for the Study of Pain (IASP) emphasizes the necessity for objective pain assessment tools to complement subjective reporting, particularly in populations unable to communicate effectively (IASP, 2020). This study contributes to this objective by providing a statistically significant method for pain classification, which could enhance diagnostic accuracy and treatment personalization. Furthermore, the differentiation of pain types—neuropathic, nociceptive, and nociplastic—remains a critical challenge in clinical settings (Treede, 2015). EEG-based classifiers hold promise in distinguishing these pain modalities by identifying specific neural signatures associated with each type. For instance, neuropathic pain may exhibit distinct connectivity patterns and oscillatory changes compared to nociceptive pain, as suggested by recent neuroimaging studies (Napadow,2010). Accurate classification can inform targeted therapeutic interventions, thereby improving patient outcomes.

The development of objective pain measurement technologies, such as the EEG-based classifier presented in this study, introduces significant ethical and medico-legal implications. The IASP underscores the importance of ensuring that such technologies are used to enhance patient care without infringing on patient autonomy or privacy (IASP, 2020). Additionally, the medico-legal landscape may be influenced by the ability to provide empirical evidence of pain states in contexts such as disability claims or legal disputes, necessitating rigorous validation and standardization of these tools to ensure their reliability and fairness (Gaskin, 2011).

While the results are promising, further research is essential to refine EEG-based pain classifiers. Expanding the training dataset to include a more diverse population and various pain etiologies will enhance the generalizability of the model. Integrating EEG data with other physiological markers, such as heart rate variability or biomarkers, could improve classification accuracy and provide a more comprehensive assessment of pain (Aguirre, 2015). Moreover, longitudinal studies are needed to evaluate the classifier’s performance over time and its ability to monitor changes in pain states in response to treatment. Addressing these areas will be pivotal in advancing EEG-based technologies toward clinical implementation, ultimately contributing to more effective and personalized pain management strategies.

## 5. Conclusion

This study provides compelling evidence for the feasibility of using ML algorithms with qEEG data to objectively classify chronic pain states. By aligning the identified EEG features with established neurological patterns in chronic pain, the classifier exhibits potential for clinical application. However, addressing ethical considerations and further validating the model across diverse populations and pain types are crucial steps toward realizing the full clinical utility of this technology.

## Data Availability

All data produced in the present study are available upon reasonable request to the authors

## Acknowledgements

Research reported in this publication was supported by the National Institute on Drug Abuse of the National Institutes of Health under Award Number R44DA046964. The content is solely the responsibility of the authors and does not necessarily represent the official views of the National Institutes of Health. The author(s) declared the following potential conflicts of interest with respect to the research, authorship, and/or publication of this article: Frank Minella is the owner of PainQx and is an investor on patents licensed by PainQx from NYU School of Medicine. Jonathan Miller, Skylar Jacobs, William Koppes, Federica Porta, and Joseph A. Lovelace were employed by PainQx at the time of the research. Fletcher White consulted on the project and was an editor of the paper. Mark Hutchinson consulted on the data analysis, interpretation and communication and was a writer and editor of the paper.

## Notes

### Summary of Updates

The manuscript has been substantially revised to improve clarity, methodological transparency, and overall scientific rigor. Key updates include a more detailed description of the study population, inclusion/exclusion criteria, and data processing pipeline. The revised version provides clearer explanations of how EEG data were prepared and analyzed, ensuring the approach is both reproducible and aligned with current best practices. The machine learning methodology has also been expanded, with a more thorough explanation of algorithm selection, model validation, and performance evaluation. Classifier metrics have been updated to include accuracy, sensitivity, specificity, and predictive values, using both development and independent test datasets. Additional updates highlight the EEG features most relevant to pain classification and contextualize their importance based on prior research. The discussion and conclusion sections have been revised to better address the clinical significance of the findings, ethical considerations, and areas for future research. Finally, Mark R. Hutchinson has been included as an author of the manuscript.

